# Artificial intelligence for ventricular arrhythmia capability using ambulatory electrocardiograms

**DOI:** 10.1101/2023.12.18.23300017

**Authors:** Joseph Barker, Xin Li, Ahmed Kotb, Akash Mavilakandy, Ibrahim Antoun, Chokanan Thaitirarot, Ivelin Koev, Sharon Man, Fernando S Schlindwein, Harshil Dhutia, Shui Hao Chin, Ivan Tyukin, William B Nicolson, G Andre Ng

**Affiliations:** Department of Cardiovascular Sciences, University of Leicester, Leicester, UK; Cardiology Department, Glenfield Hospital, University Hospitals of Leicester, Leicester, UK; Cardiology Department, Kettering Hospital, University Hospitals of Northamptonshire NHS Group, Northampton, UK; National Institute for Health Research Leicester Biomedical Research Centre, Leicester, UK; National Heart and Lung Institute, Imperial College London, UK; School of Engineering, University of Leicester, Leicester, UK; Department of Mathematics, King’s College London, London, UK

**Keywords:** Ventricular arrythmia, deep learning, risk stratification, artificial intelligence, implantable cardioverter defibrillator, neural network

## Abstract

**Question:** Can artificial intelligence (AI) be used to predict if a person is at risk of a lethal heart rhythm, based solely on an electrocardiogram (an electrical heart tracing)?

**Findings:** In a study of 270 adults (of which 159 had lethal arrhythmias), the AI was correct in 4 out of every 5 cases. If the AI said a person was at risk, the risk of lethal event was three times higher than normal adults.

**Meaning:** In this study, the AI performed better than current medical guidelines. The AI was able accurately determine the risk of lethal arrhythmia from standard heart tracings over a year away which is a conceptual shift in what an AI model can see and predict. This method shows promise in better allocating implantable shock box pacemakers (ICDs) that saves lives.

**Scientific Abstract:** *Aim:* Current clinical practice guidelines for implantable cardioverter defibrillators (ICDs) are insufficiently accurate for ventricular arrhythmia (VA) risk stratification leading to significant morbidity and mortality. Artificial intelligence offers novel risk stratification lens through which VA capability can be determined from electrocardiogram in normal sinus rhythm. The aim was to develop and test a deep neural network for VA risk stratification using routinely collected ambulatory electrocardiograms.

*Methods:* A multicentre case-control study was undertaken to assess VA-ResNet-50, our open source ResNet-50 based deep neural network. VA-ResNet-50 was designed to read pyramid samples of 3-lead 24-hour ambulatory electrocardiograms to decide if a heart is capable of VA based on the electrocardiogram alone. Consecutive adults with VA from East Midlands, UK, who had ambulatory electrocardiograms as part of their NHS care between 2014 and 2022 were recruited and compared to all comer ambulatory electrocardiograms without VA.

*Results:* Of 270 patients, 159 heterogeneous patients had a composite VA outcome. The mean time difference between the electrocardiogram and VA was 1.6 years (⅓ ambulatory electrocardiogram before VA). The deep neural network was able to classify electrocardiograms for VA capability with an accuracy of 0.76 (CI 95% 0.66 - 0.87), F1 score of 0.79 (0.67 - 0.90), AUC of 0.8 (0.67 - 0.91) and RR of 2.87 (1.41 - 5.81).

*Conclusion:* Ambulatory electrocardiograms confer risk signals for VA risk stratification when analysed using VA-ResNet-50. *Pyramid sampling* from the ambulatory electrocardiograms is hypothesised to capture autonomic activity. We encourage groups to build on this open-source model.

## Introduction

Ventricular arrhythmias (VAs) can be lethal with survival determined by access to effective defibrillation. Delays in defibrillation are associated with functional disability, care dependency and death (1). External defibrillators are not accessed in 90% of cases and current implantable cardioverter-defibrillators (ICDs) guidelines are insufficiently accurate, including fewer than 20% of all VAs (2,3). Novel artificial intelligence (AI) prediction methodologies may improve current guidelines to determine VA capability and therefore assign ICDs more accurately (4). The aim is to develop a deep neural network for VA risk stratification using routinely collected ambulatory electrocardiograms in order to predict VA capability from a patient’s normal cardiac rhythm.

## Methods

A multicentre retrospective deep-learning based case-control study was undertaken at University Hospitals of Leicester (UHL: Leicester General Hospital, Glenfield Hospital & Leicester Royal Infirmary) and University Hospitals of Northamptonshire NHS Group (UHN: Kettering Hospital), UK. Eligibility for ventricular arrhythmia cohort was consecutive adults with International Classification of Diseases 10 (ICD10) diagnoses of I47.2 ventricular tachycardia (VT) & I49.0 ventricular fibrillation/flutter (VF) with ambulatory electrocardiograms between 2014-2022. For patients with multiple ambulatory electrocardiograms, only the earliest was taken. There is no established power factor for AI studies, therefore the largest possible cohort was sought. Eligibility for the comparator cohort was consecutive adults with ambulatory electrocardiograms over a 5-day period in Nov 2022.

### Signal processing

Three-lead ambulatory 24-hour electrocardiograms (Spacelabs Lifecard CF Holter monitors 128hz) were exported from Spacelabs in ISHNE (.ecg) format. Electrocardiograms were pre-processed using a second-order Bessel filter, passband between 0.1 Hz and 50 Hz, and a notch filter at 50 Hz. R peaks in the electrocardiogram data were detected using the Pan-Tompkins algorithm. Smoothed RR intervals were obtained by a moving average filter with a factor of 40 samples before converting to HR. *Pyramid sampling* was undertaken by categorising computed heart rates into 100 distinct levels ranging from slow to fast rates; these levels were defined by uniformly distributed percentiles regardless of time of the day. One 10 second segment was selected randomly from each level and formed one input sample. This selection process was repeated 100 times for each patient and for each of the three electrocardiogram leads. As a result, for each patient, 100 3D tensors were generated in the format of 100X1280X3, (figure1).

### Data Flow

A patient-wise partitioning strategy was employed, allocating 80% for training and 20% for testing. Within the training data, we instituted a 5-fold cross-validation procedure partitioned by patients. Consequently, this yielded five distinct models from which an ensemble approach converged the predictions to deliver the final prediction model on the unseen test data. Figure 1.

**Figure 1.**
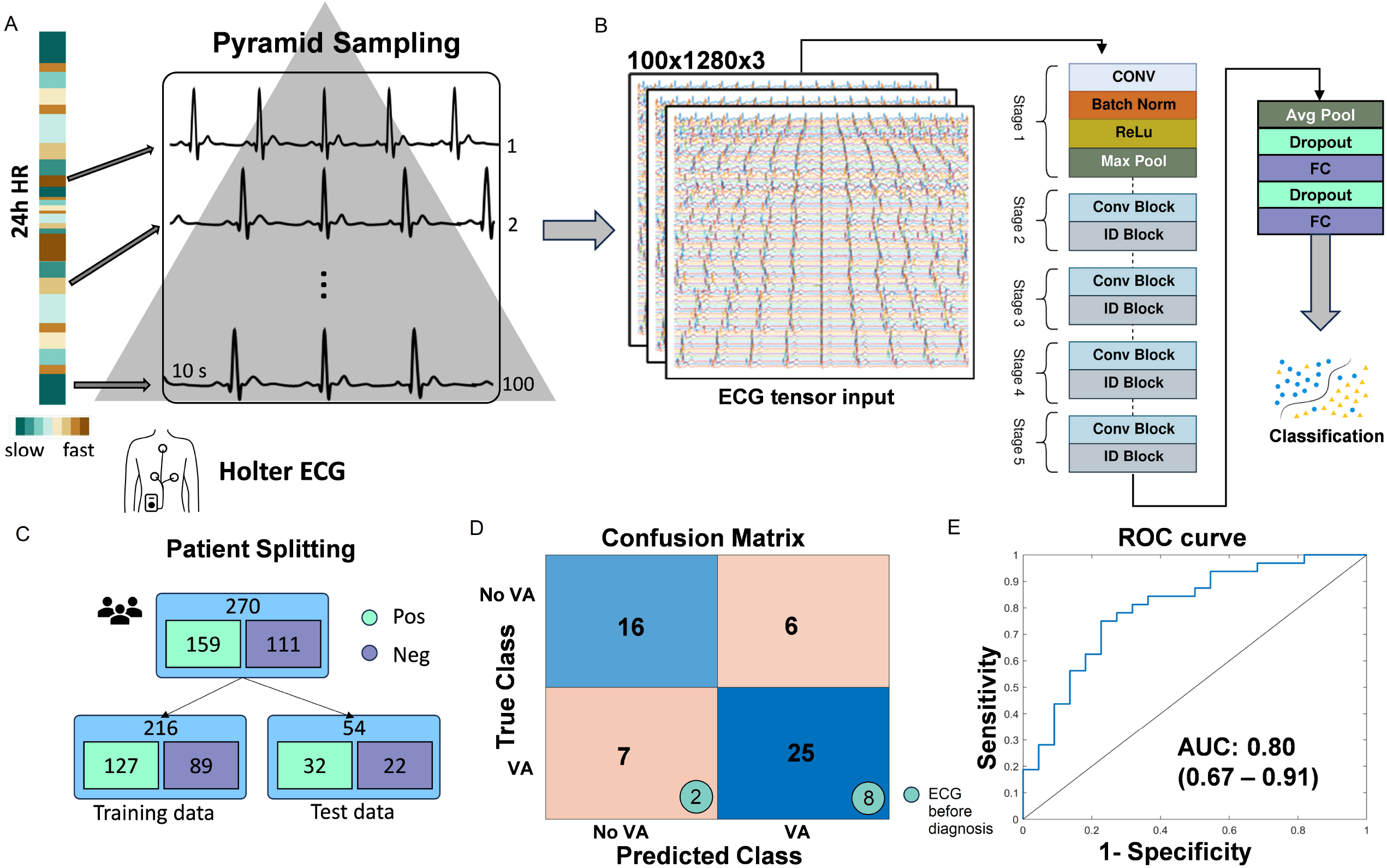
**A – Pyramid sampling schematic demonstrating 100 samples at various heart rates over 24-hour period. B - VA-ResNet-50 Architecture. C – Patient flow. D Confusion Matrix-including participants before and after ECG. E – Receiver Operator Characteristic Curve. Abbreviations: Receiver Operator Characteristic (ROC), ventricular arrhythmia (VA), electrocardiogram (ECG), average pooling (avg pool), batch normalisation (batch norm), identity block (ID block), convolutional block (conv block), fully connected (FC), Rectified Linear Unit (ReLu).**

### Deep learning architecture

A transfer-learned, customised ResNet-50 based convolutional neural network was employed with a modified input layer we name: VA-ResNet-50. The architecture was organized into blocks of convolutional and identity layers with 53 convolutional layers using various filter sizes, 53 batch normalisation layers, 49 ReLU layers, and 5 pooling layers. In addition to these, we have incorporated 3 fully connected layers, two dropout layers, L2 regularization (regularization factor of 0.01), a softmax layer, and a classification layer, (figure 1). We employed a learning rate of 1e-4, a decay factor of 0.1 every 5 epochs, for 20 epoch max with early stopping at 2 consecutive epochs and adam optimisation. The batch size was 64, the dropout layer rate was 0.6. Open-source model is available on Github (5). The model served to take 100x3D ECG tensors per patient that are individually classified by the model to generate the averaged class probability score for the per-patient prediction with a threshold arbitrarily set to 0.5.

### Statistical analysis

The performance of the neural network was evaluated in the test set using metrics such as overall accuracy, F1 score, sensitivity, specificity, AUC, AUPCR. Confidence intervals for performance metrics were calculated using bootstrapping with 1000 iterations. All analyses were completed in MATLAB (Mathworks, USA).

### Permissions and reporting

Study permissions were granted from the respective institutional review committees; registration numbers; UHN: REF8882 & UHL: REF11434. This paper was reported according to STARD2015 reporting guidelines.

## Results

The study comprised 270 patients - 178 UHN and 92 UHL patients. VA occurred in 159 patients (mean [CI:95%] age 61 years [57-65]; 78 female (49%)) compared to 111 patients without VA (age 58 years [54-62]; 56 female (51%). The VA positive and negative cohorts differed in proportions of cardiovascular risk factors, electrocardiographically distinct diagnoses and cardiomyopathies but all diagnoses were represented within both cohorts. For the composite outcome of VA, VT was 88% (n=140) whilst 24% (n=39) represented VF and 6% represented both VT & VF diagnoses, (table 1). The mean time difference between the electrocardiogram and VA was 1.6 years with 27% of the cohort having an ambulatory electrocardiogram before the VA.

**Table 1:** Patient characteristics & performance metrics. Abbreviations: area under the receiver operator curve (AUC), area under the precision-recall curve (AUCPR), confidence interval (CI), positive predictive value (PPV), negative predictive value (NPV), relative risk (RR), ventricular arrhythmia (VA), electrocardiogram (ECG).

| Patient characteristics | Ventricular arrhythmia (n=159) | No ventricular arrhythmia (n= 111) |
| --- | --- | --- |
| **Demographics** |  |  |
| Age in years n (CI 95%) | 61 (57-65) | 58 (54-62) |
| Female n (%) | 78 (49%) | 56 (51%) |
| **Cardiomyopathies** |  |  |
| Ischaemic heart disease n (%) | 92 (59%) | 19 (17%) |
| Inherited Cardiomyopathy n (%) | 24 (15%) | 3 (3%) |
| Hypertrophic cardiomyopathy n (%) | 7 (4%) | 1 (1%) |
| Heart failure (any) n (%) | 47 (30%) | 13 (12%) |
| Dilated Cardiomyopathy n (%) | 10 (6%) | 1 (<1%) |
| Myocarditis n (%) | 2 (1%) | 0 (0%) |
| Valvular Heart Disease n (%) | 69 (43%) | 11 (10%) |
| **Cardiovascular risk factors** |  |  |
| Hypertension n (%) | 98 (61%) | 34 (31%) |
| Type 2 diabetes n (%) | 16 (10%) | 5 (5%) |
| Chronic obstructive pulmonary disease n (%) | 22 (14%) | 6 (5%) |
| Chronic Kidney Disease n (%) | 1 (<1%) | 1 (<1%) |
| Dyslipidaemia n (%) | 51 (32%) | 16 (14%) |
| Syncope n (%) | 22 (14%) | 7 (6%) |

**Table 1: Patient characteristics & performance metrics.**
| Electrocardiographically distinct diagnoses |  |  |
| --- | --- | --- |
| Atrial fibrillation or flutter n (%) | 56 (35%) | 14 (13%) |
| Left bundle branch block n (%) | 18 (11%) | 3 (3%) |
| Complete Heart Block n (%) | 3 (2%) | 0 (0%) |
| Ventricular arrhythmia outcomes |  |  |
| Ventricular tachycardia n (%) | 131 (82%) | 0 (0%) |
| Ventricular fibrillation/ flutter n (%) | 36 (23%) | 0 (0%) |
| Both ventricular tachycardia & fibrillation/ flutter | 8 (5%) | 0 (0%) |
| Presence of implantable cardioverter defibrillator | 23 (15%) | 0 (0%) |
| Time between ambulatory electrocardiogram and ventricular arrhythmia |  |  |
| Mean time difference | 1.6 years (n=159) | NA |
| ECG collected before VA | 0.4 years (n=43, 27%) | NA |
| ECG collected after VA | 2.1 years (N=117, 73%) | NA |
| VA-ResNet-50 performance |  |  |
| Performance metric | Train set - 80% - 5-fold cross validation results | Test set - 20% - ensemble per patient results |
| Accuracy (CI 95%) | 0.71 (0.6 - 0.82) | 0.76 (0.66 - 0.87) |
| AUC (CI 95%) | 0.76 (0.64 - 0.88) | 0.80 (0.67 - 0.91) |
| AUCPR (CI 95%) | 0.77 (0.65 - 0.89) | 0.81 (0.64 - 0.91) |
| F1 Score (CI 95%) | 0.76 (0.66 - 0.86) | 0.79 (0.67 - 0.90) |
| Balanced accuracy (CI 95%) | 0.70 (0.53 - 0.83) | 0.76 (0.64 - 0.87) |
| PPV (CI 95%) | 0.75 (0.65-0.85) | 0.81 (0.67 - 0.95) |
| NPV (CI 95%) | 0.67 (0.53 -0.81) | 0.70 ( 0.51 - 0.89) |
| RR (CI 95%) | 2.32 (1.36 - 3.28) | 2.87 (1.41 - 5.81) |
| Sensitivity (CI 95%) | 0.62 (0.48 - 0.76) | 0.78 (0.64 - 0.92) |
| Specificity (CI 95%) | 0.78 (0.67 - 0.89) | 0.73 (0.52 - 0.91) |

### Deep learning performance

For the testing dataset, from sinus rhythm the model was able to classify by VA capability with an F1 score of 0.79 (0.67 - 0.90). Figure 1 displays the confusion matrix. The accuracy was 0.76 (CI 95% 0.66 - 0.87). Figure 1 shows the ROC curve for the test dataset, the AUC was 0.8 (0.67 - 0.91). The relative risk was 2.87 (1.41 - 5.81), (table 1).

## Discussion

VA-ResNet-50, a deep neural network classifier for ambulatory electrocardiograms, demonstrates signals exist for VA capability from normal intrinsic cardiac rhythm. This re-look at ubiquitously available, non-invasive, cheap cardiovascular patient data holds promise to assign ICDs with greater precision and more economically than current practice (6). The AUC of 0.8 is consistent with a growing body of evidence for VA prediction with AI (4,10). Specifically for ambulatory electrocardiograms we report lower AUC than unpublished results of Fiorena *et al*. (AUC=0.91) to predict incident sustained VT (7). This difference is likely because of time disparities between electrocardiogram and VA between cohorts, 2 weeks vs. 1.6 years. This work improves on work published by Sammani *et al*. who describe an AUC of 0.67, albeit with an explainable autoencoder model within a cohort of dilated cardiomyopathy (8). This difference might be explained by their refined “life-threatening” VA outcome - granularity of outcome not available to ICD10. Our newly termed *pyramid sampling* could explain our good performance as autonomic nervous activity is a prognostic marker for VAs which manifests over a range of heart rates (9).

## Limitations

Retrospective recruitment of VA meant ambulatory electrocardiograms were frequently available after VA which introduces survivor bias. The traumatic nature of VA may induce electrophysiological changes to be detected. The heterogeneity of the cohort is representative of real-world VA but precludes the convention of mechanistic understanding required of evidence-based medicine. The comparator cohort is similarly comorbid but healthier than the original intended comparator cohort meaning the model is at risk of classifying heart health index as opposed to a VA risk specifically. The intended comparator cohort; those with ICDs and no VA; were not available because they do not undergo ambulatory electrocardiograms due device EGM availability. The outcomes were derived from secondary care billing data only which can result in misclassification. The ad-hoc data collection strategy likely confounds external validity, though this recruitment strategy is consistent with the field due to the sudden and unexpected nature of VA (4). Our current analysis does not include a systematic assessment of model stability to random and adversarial perturbations to data and model structure (10). This, however, will be the topic of our future work.

## Conclusion

Normal sinus ambulatory electrocardiograms contain signals for VA risk stratification when analysed using our open-source VA-ResNet-50 deep neural network. *Pyramid sampling* from the ambulatory electrocardiograms is hypothesised to capture autonomic activity. Importantly VA-ResNet-50 is agnostic of cardiomyopathy. The retrospective, *ad hoc* recruitment strategy limits generalisability. Prospective validation within other cohorts is planned and we encourage other groups to build on this open-source model.

## Data Availability

All data produced in the present study are available upon reasonable request to the authors.

https://github.com/lixin0121/VA-resnet-50

## Acknowledgements

We acknowledge the guidance and expertise of Ms Michelle Newton, Mr Alan Bruce, Mr Graham Williams, Mr Richard Weatherhead, Ms Louise Waller, Mr Adam Brown, Ms Sally Berrill, Ms Kathy Clarke and Ms Mei-Mei Chueng for their assistance in navigating the permissions and respective data infrastructures of the involved institutions. We acknowledge the cardiac physiologist expertise of Ms Jen Hambleton, Mr Vasco Rodrigues and Ms Vanisha Jonas in data acquisition and analyses.

